# Blood pressure variability and night-time dipping assessed by 24-hour ambulatory monitoring: cross-sectional association with cardiac structure in adolescents

**DOI:** 10.1101/2020.11.20.20235473

**Authors:** Lucy Goudswaard, Sean Harrison, Daniel Van De Klee, Nishi Chaturvedi, Debbie Lawlor, George Davey Smith, Alun Hughes, Laura D Howe

## Abstract

Greater blood pressure (BP) variability and reduced night-time BP dipping are associated with cardiovascular disease risk independently of mean BP in adults. This study examines whether these associations are apparent in a general population of adolescents. A cross-sectional analysis was undertaken in 587 UK adolescents (mean age 17.7 years; 43.1% male). BP was measured in a research clinic and using 24-hour ambulatory monitoring. We examined associations (for both systolic and diastolic BP) of: 1) clinic and 24-hour mean BP; 2) measures of 24-hour BP variability: standard deviation weighted for day/night (SDdn), variability independent of the mean (VIM) and average real variability (ARV); and 3) night-time dipping with cardiac structures. Cardiac structures were assessed by echocardiography: 1) left ventricular mass indexed to height^2.7^ (LVMi^2.7^); 2) relative wall thickness (RWT); 3) left atrial diameter indexed to height (LADi) and 4) left ventricular internal diameter in diastole (LVIDD). Higher systolic BP was associated with greater LVMi^2.7^. Systolic and diastolic BP were associated with greater RWT. Associations were inconsistent for LADi and LVIDD. There was evidence for associations between both greater SDdn and ARV and higher RWT (per 1 SD higher diastolic ARV, mean difference in RWT was 0.13 SDs, 95% CI 0.045 to 0.21); these associations with RWT remained after adjustment for mean BP. There was no consistent evidence of associations between night-time dipping and cardiac structure. In this general adolescent population study, associations between BP variability and cardiac structure were apparent. Measurement of BP variability might benefit cardiovascular risk assessment in adolescents.

## Introduction

Higher blood pressure (BP) is associated with an increased risk of cardiovascular disease (CVD) [1]. However, BP is inherently variable, and under a typical circadian rhythm night-time BP is lower than daytime [2]. Loss of this nocturnal dipping pattern in the general population of adults has been shown to be associated with cardiovascular events and all-cause mortality, independent of 24-hour BP [2, 3]. There is also evidence that non-circadian variability in BP may be associated with cardiovascular disease [2, 4, 5].

Cardiovascular pathology starts in early life, with childhood BP levels known to track across life [6], and early adulthood BP relating to mortality from CVD [7]. In adults, higher left ventricular (LV) mass and left atrial enlargement are both associated with higher risk of CVD [8, 9] and are considered evidence of target organ damage [10]. Another measure of left heart function, relative wall thickness (RWT, a measure of remodelling [11]), has been suggested to be predictive of stroke among adult populations [12, 13]. We previously demonstrated that in 17 year-olds that higher body mass index (BMI) is causally related to higher LV mass indexed to height^2.7^ (LVMi^2.7^) [14], suggesting that there is meaningful variation in cardiac structure measures in early adulthood. A study in adults from the general population indicated a positive association between BP variability and LVMi [15]. Associations between BP variability and cardiac structures in children with suspected hypertension have been explored [16], but it is unclear if any associations are apparent in a general population of adolescents.

In this study, we used data from a prospective cohort study of 587 UK adolescents to assess the cross-sectional associations of mean BP (from clinic measurements and ambulatory monitoring), BP variability, and night-time dipping, with measures of cardiac structure at age 17, determined by echocardiography. The measures of cardiac structure we consider are 1) LV mass (LVM), 2) RWT [11], 3) left atrial diameter (LAD), and 4) left ventricular internal diameter during diastole (LVIDD, a measure of the initial stretching of cardiomyocytes before contraction (preload)) [17]. Together these represent a comprehensive assessment of left heart structure, with functional significance [18].

## Methods

### Participants

ALSPAC is a population-based birth cohort. The study recruited pregnant women from the Avon area (Bristol) in the South West of England, with an expected delivery date between 1^st^April 1991 and 31^st^ December 1992 [19]. From the 15,643 pregnant women enrolled, 14,889 children were born and alive at one year [19, 20] (Figure 1). Since birth, participants have been followed up, using questionnaires, links to routine data, and research clinics. The study website provides further details of the cohort and a data dictionary http://www.bris.ac.uk/alspac/researchers/data-access/data-dictionary/. Approval was obtained from the local ethics committee and the ALSPAC Law and Ethics committee.

**Figure 1:**
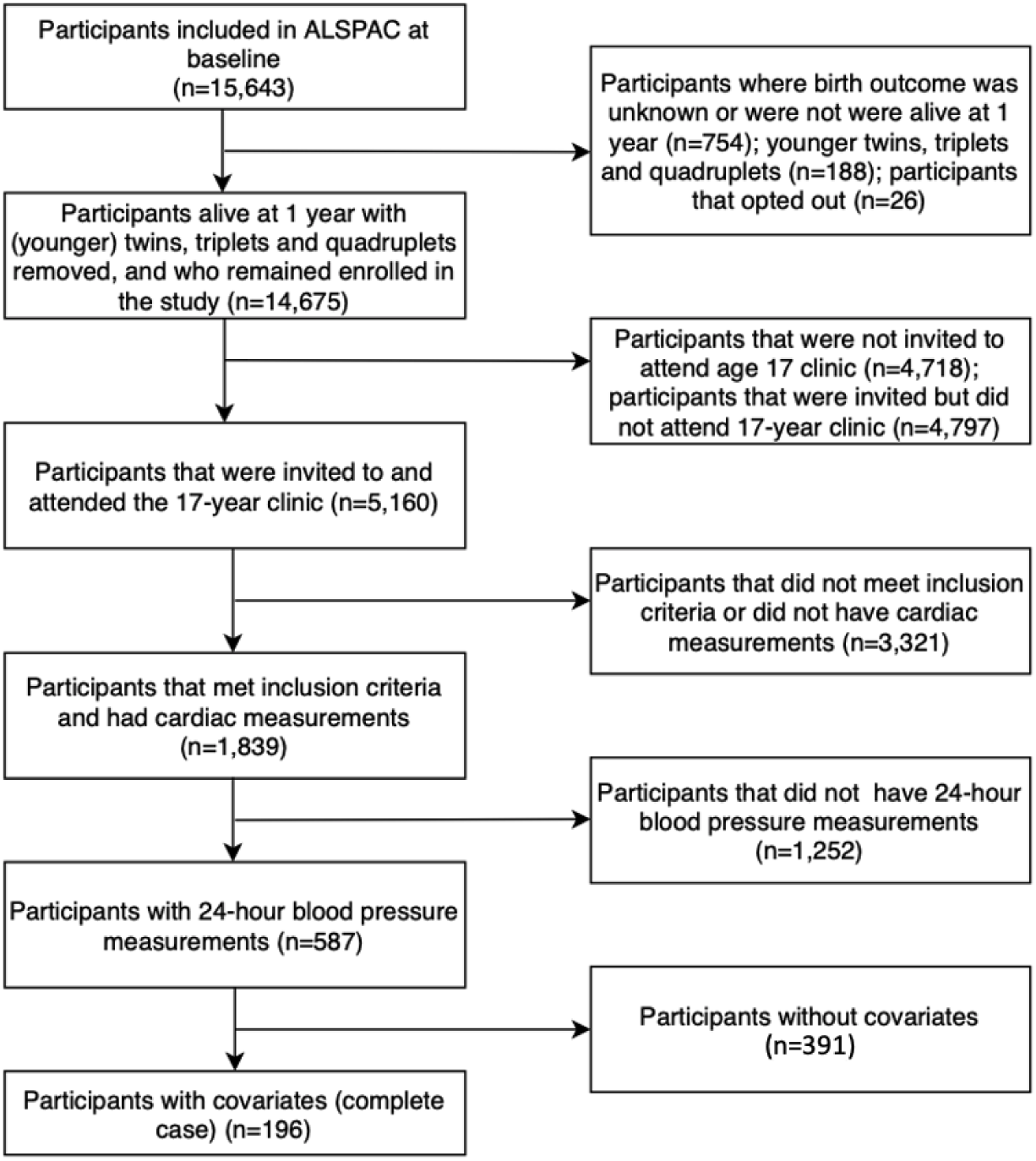
a STROBE diagram detailing how the study cohort was selected from the baseline Avon Longitudinal Study of Parents And Children (ALSPAC) participants.

### Inclusion/Exclusion Criteria

This was a cross-sectional study conducted in participants who attended the 17-year follow-up clinic of ALSPAC. Participants were eligible if they attended both the echocardiography and the 24-hour blood pressure sub-studies at the 17-year clinic visit. We a priori decided to exclude participants if they were pregnant or reported taking antihypertensive medication or having a congenital cardiac anomaly, but this did not apply to any participants in the study.

### Exposures

#### 1) Clinic and 24-hour ambulatory blood pressure measurements

Clinic systolic blood pressure (SBP) and diastolic blood pressure (DBP) were measured with an OMRON 705 IT oscillometric BP monitor (Omron Corporation, Kyoto, Japan) with the participant sitting and at rest with their arm supported. Readings were taken in accordance with European Society of Hypertension guidelines [21]. We used the average of the final two measures from the right arm in our analyses.

Participants were fitted with a 24-hour ambulatory blood pressure monitor (ABPM) (Spacelabs 90217, Washington, U.S.) according to the manufacturer’s instructions. This measured their brachial BP, with readings taken every 30 minutes during the day and hourly at night. Participants were permitted to perform usual physical activities, although a diary of activities was recorded. Daytime and night-time were defined by the participant. The expected maximum number of total readings per participant therefore varied depending on the duration of the night-time period. For this study, we included participants with at least 14 readings during the self-defined daytime and at least 5 readings during the self-defined night-time [22, 23].

We estimated the mean 24-hour SBP and DBP using the ABPM data, and also estimated the daytime and night-time means for SBP and DBP.

#### 2) Measures of blood pressure variability

We estimated variability in the 24-hour systolic and diastolic measures in three different ways. 1) Standard deviation weighted for daytime and night-time (SDdn) [24], calculated as: 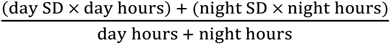. 2) Average real variability (ARV), derived as the average of the differences between consecutive BP measurements [25], using the formula: 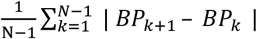, where N is the number of valid blood pressure (BP) readings and k is the number of the individual reading. To derive this variable, each individual blood pressure reading and the order of readings was required (at least 14 daytime readings and 5 night-time readings). 3) Variability independent of the mean (VIM) [26], derived using the formula: 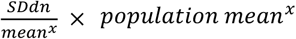, where *x* is derived from the regression coefficient β from the equation: ln(*SD*) *= α* + *β* ln(*mean*).

#### 3) Dipping variables

We estimated night-time dipping as a percentage difference between daytime and night-time means 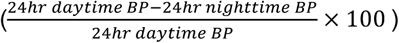 [27]. We considered participants with ≥10% reduction in night-time BP compared to daytime BP as ‘normal dippers’, and those with <10% reduction or an increase as ‘non-dippers’ in a binary dipping variable [3, 27]. We also grouped participants into four dipping groups of 1) Normal dippers (>10%, ≤20%), 2) Non-dippers (>0%, ≤10%), 3) Extreme dippers (>20%) and 4) Risers (<0%) to allow comparison of dipping distribution with previous studies [28], however only the simpler continuous and dichotomised variables are included as an exposure in our analyses because of the relatively small sample size.

### Outcomes: Echocardiography Measurements

Echocardiography was performed on a quasi-random subsample (based on date of research clinic attendance) using an HDI 5000 ultrasound machine (Philips, Massachusetts, U.S.) equipped with a P4-2 Phased Array ultrasound transducer. One of two echocardiographers examined participants using a standard examination protocol, in accordance with the American Society of Echocardiography (ASE) guidelines [29]. All measures were made in end diastole and were calculated as the mean of three measurements. LV mass was calculated from end-diastolic ventricular septal wall thickness (SWTd), left ventricular dimension (LVIDd), and left ventricular posterior wall thickness (PWT) according to the ASE formula: 0.8 × (1.04 × [(*SWT* + *LVIDD* + *PWT*)^3^ − (*LVIDD*)^3^]) + 0.6. LV mass was then indexed to height^2.7^ (LVMi^2.7^) using the Troy formula in order to account for differences in body sizes [30]. Left atrial diameter was indexed to height [9] (LADi). Relative wall thickness (RWT)was calculated using the formula 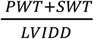.

### Confounders

We considered variables as confounders if they had plausible relations with BP and cardiovascular risk [31]. Maternal confounders were self-reported in questionnaires completed during pregnancy: educational attainment (categorised as university degree or higher, Advanced-levels (exams usually taken around 18 years and necessary for university entry), Ordinary-levels (exams usually taken around 16 years, which was the minimum UK school leaving age at the time these participants were this age), or lower than Ordinary-levels, including vocational education); pre-pregnancy body mass index (BMI; in kg/m^2^); age at delivery (categorised as <25 years, 25-35 years, and >35 years); parity, and highest head of household occupational social class. We selected these maternal variables as the mother’s socioeconomic position (SEP) represents the participant’s family SEP. SEP has been shown to influence BMI (a key determinant of both BP and LVM [14]), blood pressure [32], and left ventricular structure [33]. Maternal pre-pregnancy BMI has also been shown to affect offspring BP and cardiovascular outcomes [34].

Child-based confounders were from a combination of self-reported questionnaire and clinic-based data. These include: age (in months) at year 17 clinic visit; smoking at age 17 (<1 or ≥1 cigarette per week from self-report); minutes of moderate to vigorous physical activity at age 15 assessed by uniaxial ActiGraph accelerometer (Florida, U.S.) and used as quintiles in the analysis; percentage fat mass (assessed by dual energy-X-ray absorptiometry (DXA) at the 17-year clinic using a Lunar prodigy narrow fan beam densitometer); and height measured at the 17-year clinic using a Harpenden stadiometer (Holtain Ltd, Crymych, UK). These child-based variables were selected as they likely affect cardiovascular health [14].

### Statistical analysis

All analyses were performed using Stata version 15.1 (StataCorp, TX). We used multivariable linear regression to estimate the associations between all blood pressure exposures and cardiac structure outcomes defined above. We standardised all exposures and outcomes before analysis to have a mean of zero and SD of one. As such, all regression results are interpreted as the SD change in the outcome for a SD change in the exposure. For the binary dipping variables, the regression result can be interpreted as the change in outcome variable in SDs comparing the non-dippers category with the dippers.

Associations between each of the 18 BP exposures (for both SBP and DBP: clinic BP, 24h mean BP, mean daytime BP, mean night-time BP, SDdn, ARV, VIM and continuous and binary dipping variables) and 4 measures of cardiac structure (LVMi^2.7^, LADi, RWT, LVIDD) were assessed using multivariable linear regression. Three models were estimated: i) adjustment for sex and age at year 17 clinic visit, ii) additional adjustment for potential confounders: maternal education, age at delivery, parity, pre-pregnancy BMI; household socio-economic class; smoking at age 17; minutes of moderate to vigorous physical activity at age 15; DXA-determined fat mass and height and height^2^at age 17, iii) further adjustment for average 24-hour blood pressure (systolic or diastolic as appropriate for the exposure) to evaluate whether any associations between BP variability and dipping were independent of 24-hour average BP.

To test for interactions between sex and each exposure, we regressed each outcome on each exposure, with sex and an interaction term for the exposure and sex as covariables. There was no strong evidence of any interactions by sex from these analyses (p>0.1 for all interaction terms), and as such, all results are presented for males and females combined. To check for linearity of a) blood pressure - cardiac structure and b) mean blood pressure – blood pressure variability associations, we conducted likelihood ratio tests comparing models were fifths of the exposure variable were treated as numeric and categorical variables. There was no evidence of non-linearity in the associations between blood pressure variables and cardiac structure outcomes, and so results are presented with continuous measures of blood pressure as the exposures. As a sensitivity analysis and to account for nonlinearity in the associations between blood pressure and blood pressure variability, the association between blood pressure variability/dipping exposures and cardiac structure outcomes were also explored adjusting for categorical fifths of average blood pressure. We did not correct the results for multiple testing, as multiple testing correction emphasises the inappropriate dichotomisation of p-values into significant versus non-significant [35-38]. Furthermore, in this analysis, exposures are correlated measures of a single underlying construct BP, and outcomes are measures of a single underlying construct, cardiac structure. A Bonferroni multiple testing correction would therefore be over-conservative. We interpret the overall pattern of results rather than focusing on single p-values, and use the magnitude of coefficients and confidence intervals to assess the strength of associations.

### Missing Data

Of the 587 participants with complete data on all 18 exposures and 4 outcomes, 196 (33.3%) also had complete data including all confounders. In the full dataset, individual confounder variables were missing between 0% and 43.4% of observations, with eight of 11 variables having less than 13% missingness (Supplementary Table 1a). We used multivariate multiple imputation by chained equations to impute missing confounder data [39, 40]. The imputation model included all exposures (excluding dipping variables, which were derived from other variables in the imputation model), outcomes and confounding variables, as well as weight and BMI at age 17, and maternal height. Fully conditional specification was used, with linear regression for continuous variables, multinomial regression for categorical variables and logistic regression for binary variables (Supplementary Table 1b). We created twenty imputed datasets and used Rubin’s rules to combine analysis results. Variable distributions were consistent between the imputed and the observed data sets (Supplementary Table 1a). We also conducted a complete case sensitivity analysis in the 196 participants with complete data for all variables (Supplementary Table 2).

## Results

### Participant characteristics

A total of 587 participants were included in our analysis. Figure 1 shows how this cohort size was reached from the participants enrolled in ALSPAC at baseline. Compared with the full ALSPAC cohort, the participants included in our analysis tended to have mothers who were more educated and older when the participant was born and be from a family with a higher head of household occupational social class; females were also more likely to be included. Clinic blood pressure, minutes of moderate to vigorous physical activity at age 15 and DXA-determined fat mass were similar compared with the full ALSPAC cohort (Supplementary Table 1b).

Of the included participants, 43.1% were male, mean age was 17.7 (SD 0.3) years, 2.1% reported smoking 1 or more cigarettes a week. Mean clinic systolic and diastolic blood pressure were 114.4 mmHg (SD 9.7 mmHg) and 64.5 mmHg (SD 5.8 mmHg), respectively (table 1). 22.3% of participants were categorised as non-dippers for systolic BP, and 6.3% for diastolic BP. Mean LVMi^2.7^ was 27.7 g/m^2.7^ (SD 5.9 g/m^2.7^), LADi was 1.88 cm/m (SD 0.22 cm/m), LVIDD was 4.52 cm (SD 0.44 cm) and RWT was 0.37 (SD 0.06).

**Table 1.**
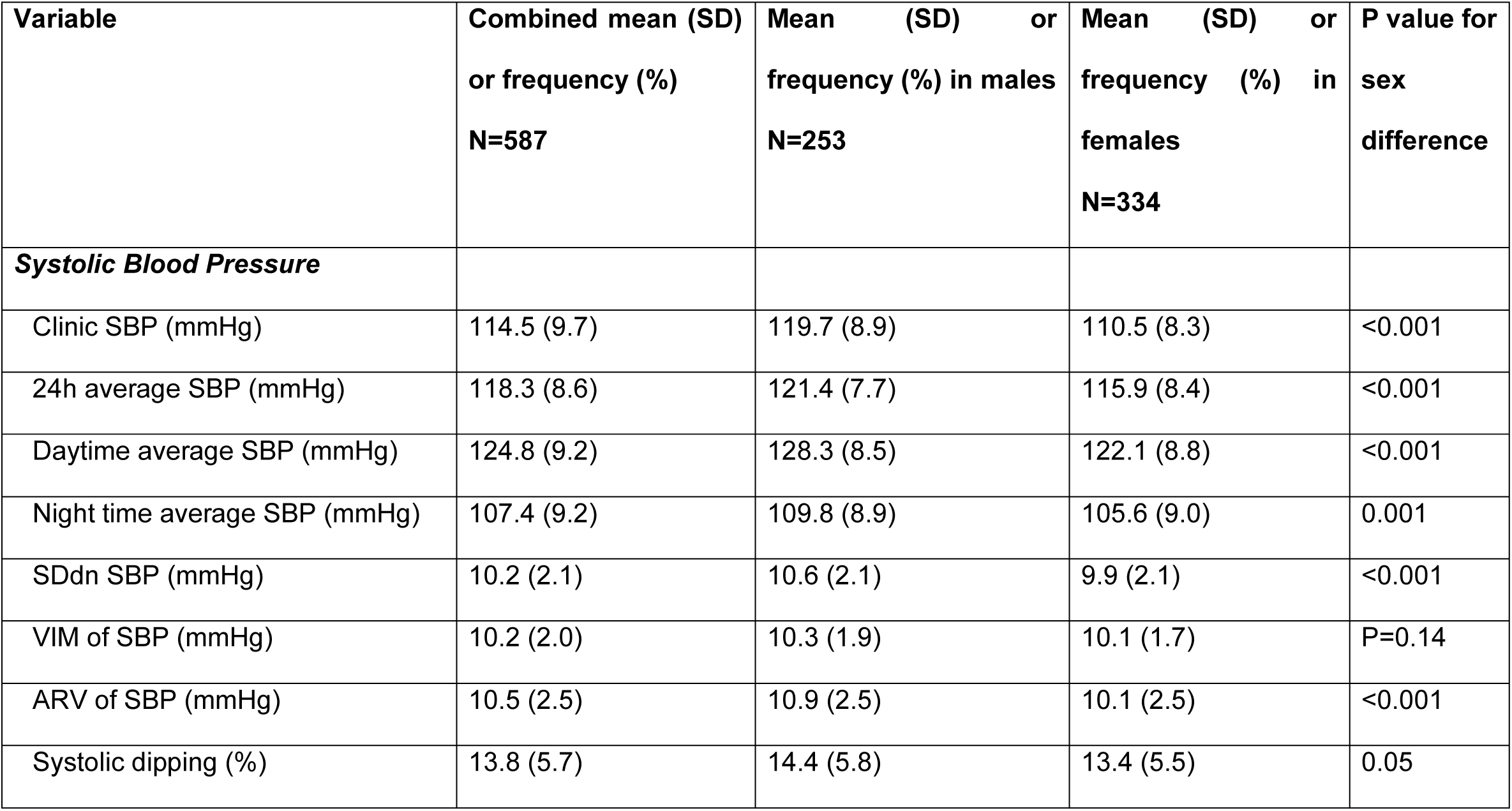
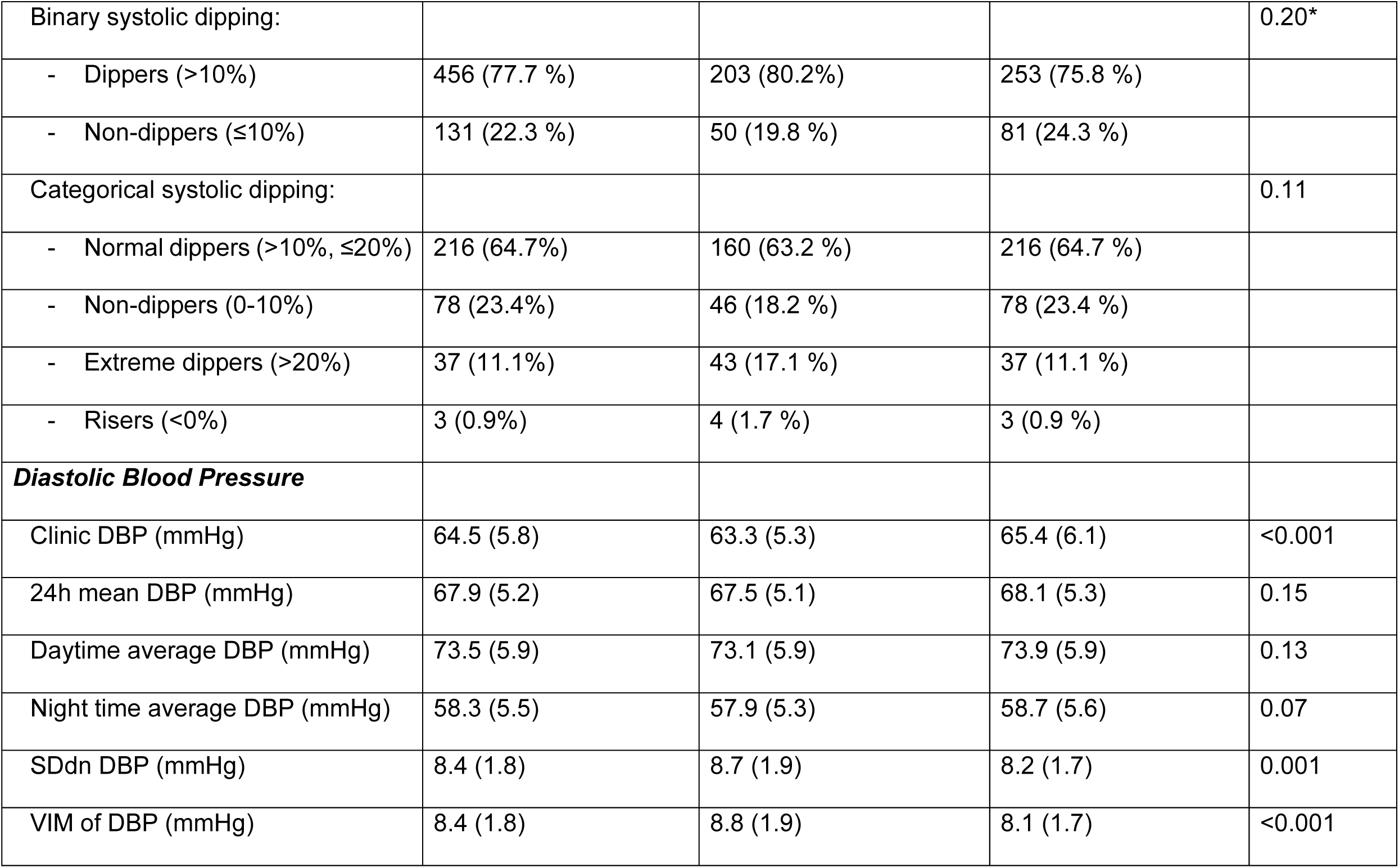
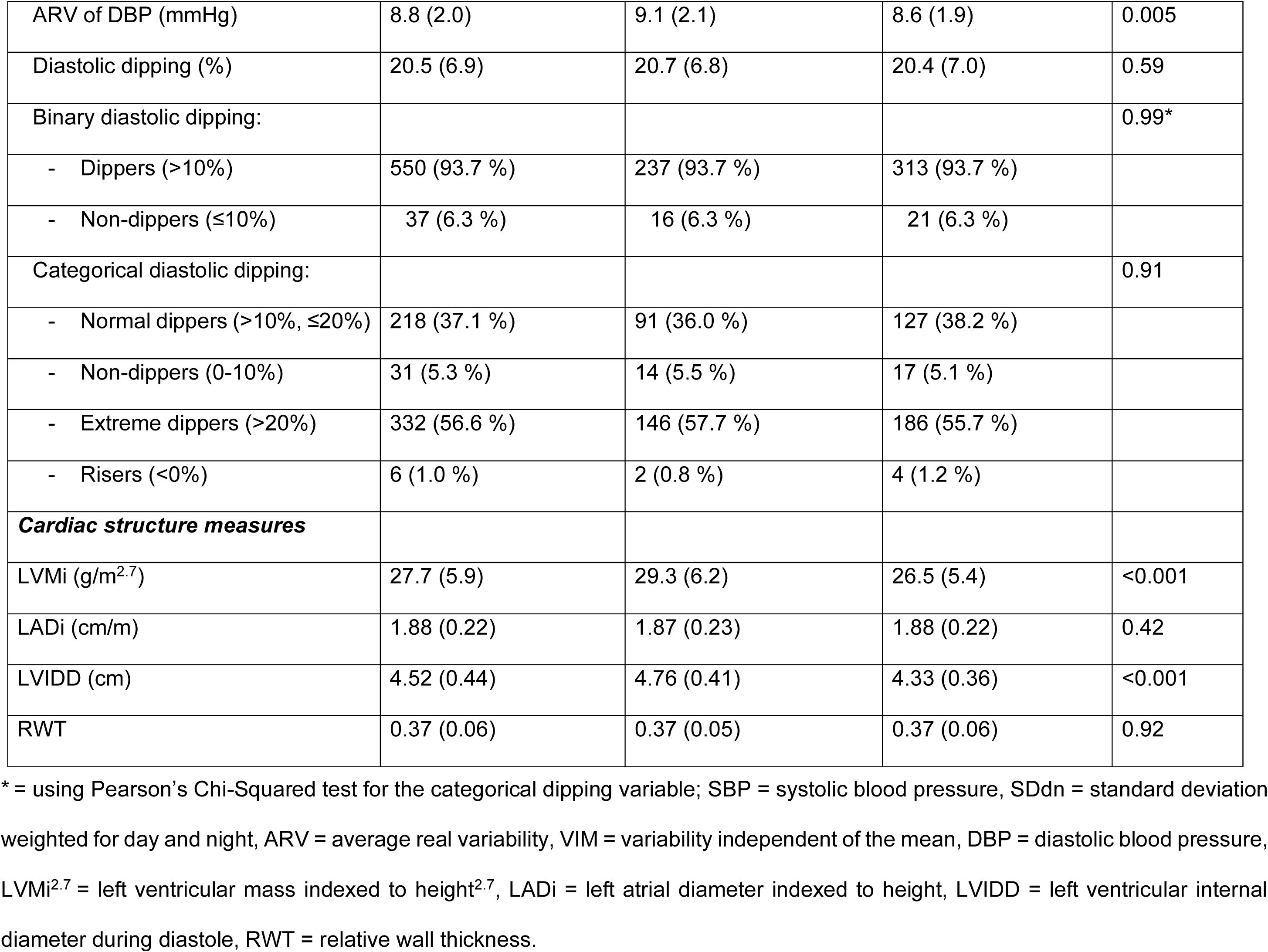
Measures of blood pressure and cardiac structure for participants included in the analysis, N=587

Males tended to have higher systolic blood pressure, pulse pressure and mean arterial pressure, while females had higher diastolic blood pressure. Night-time dipping was similar between sexes. Males tended to have higher systolic and diastolic BP variability than females. Ventricular measures were higher in males, while atrial index and wall thickness were similar between sexes (Table 1).

### 1) Associations between clinic BP measurements and cardiac structures

Clinic SBP was associated with higher LVMi^2.7^ (β = 0.23 SDs per SD increase in SBP, 95 % CI 0.15 to 0.32, P=1.6×10^−7^) and higher RWT (β = 0.29 SDs per SD increase in SBP, 95 % CI 0.19 to 0.39, P=1.2×10^−8^) after adjustment for confounders (Table 2). There was no evidence of associations with LADi or LVIDD.

**Table 2.**
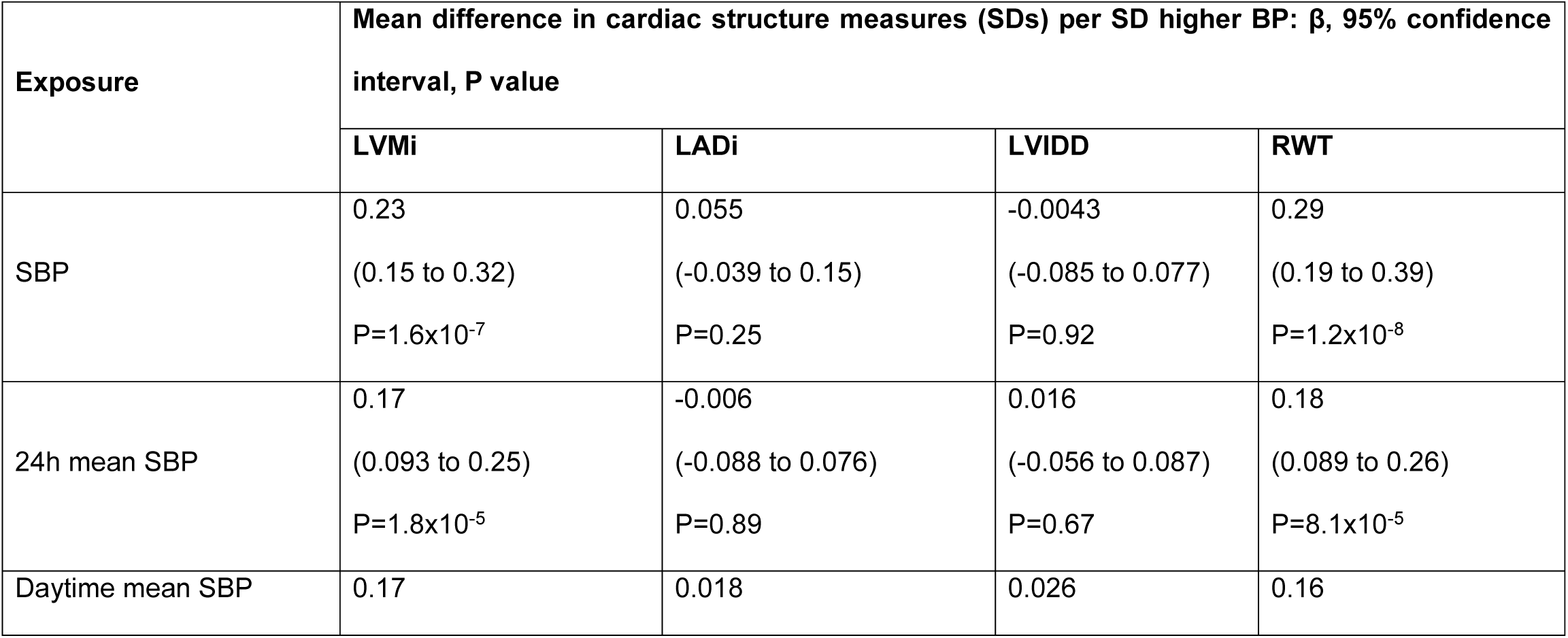
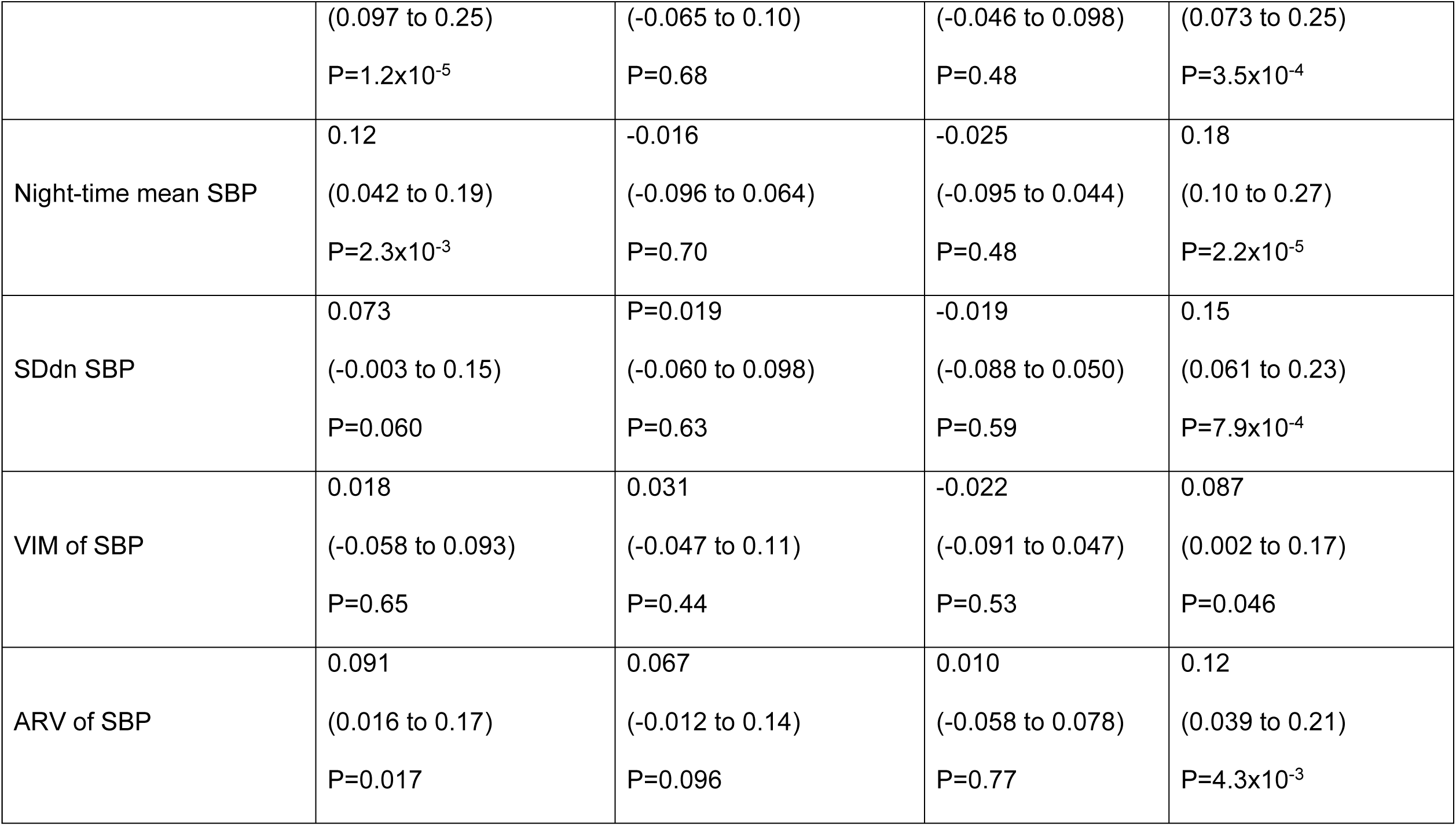
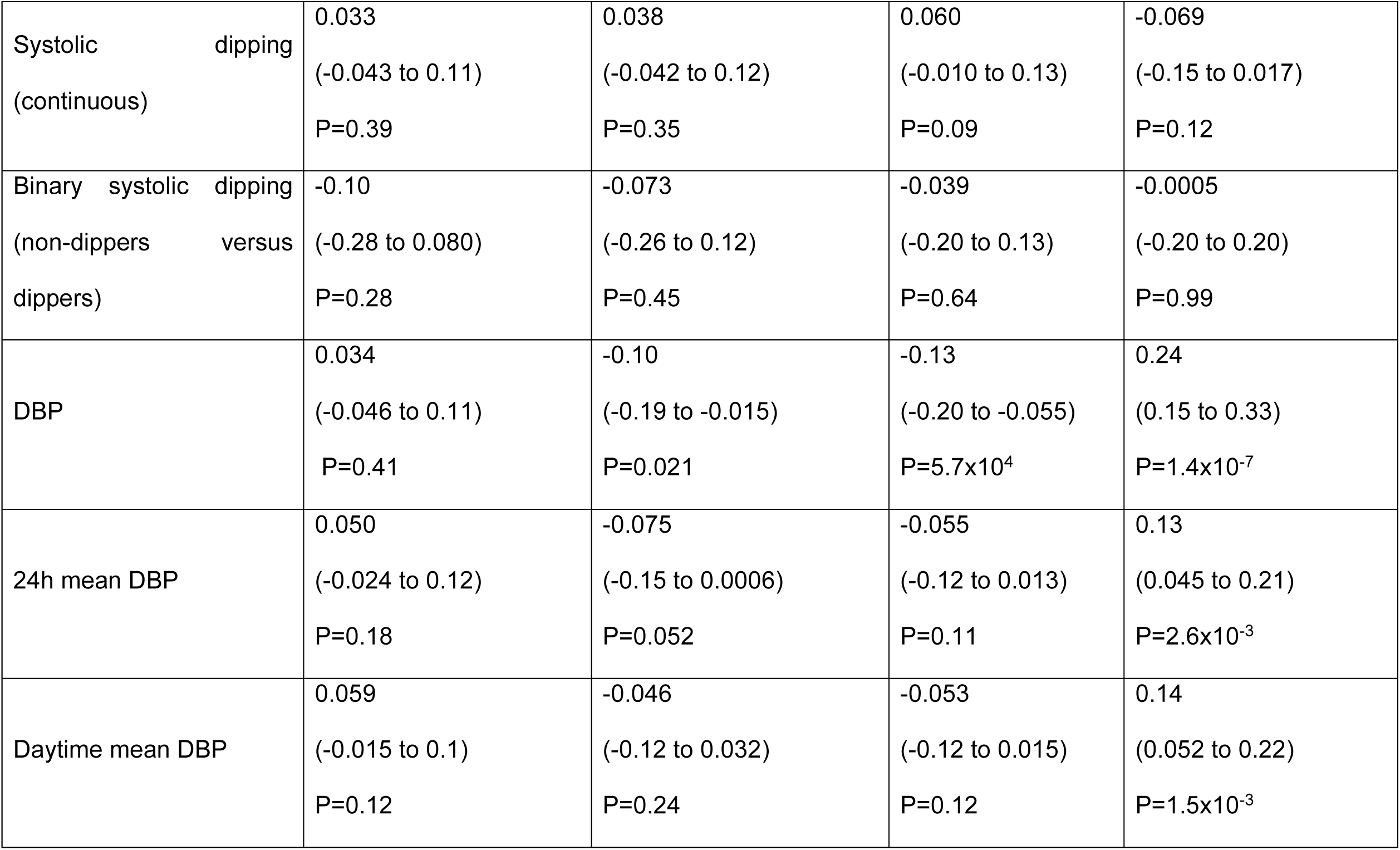
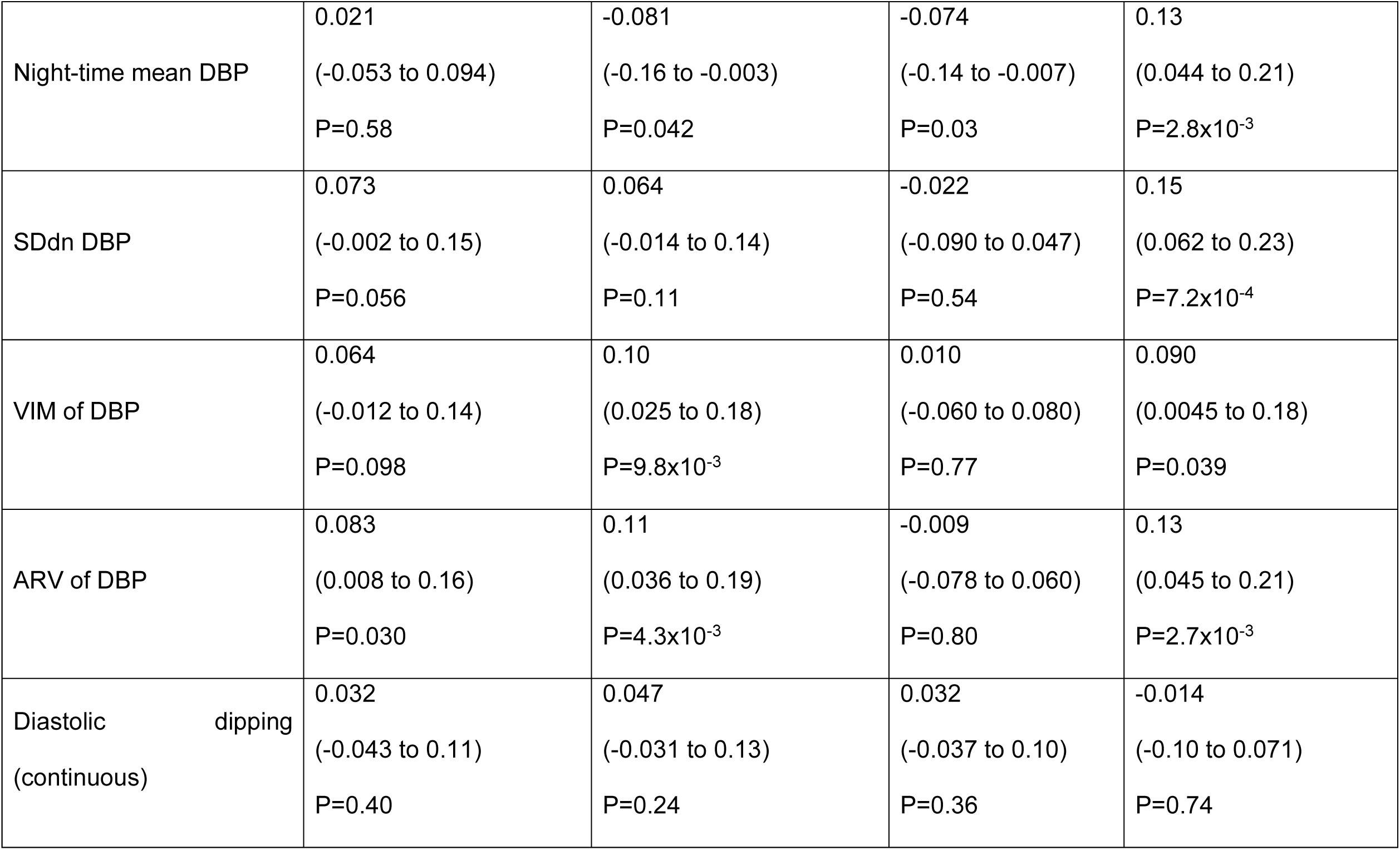
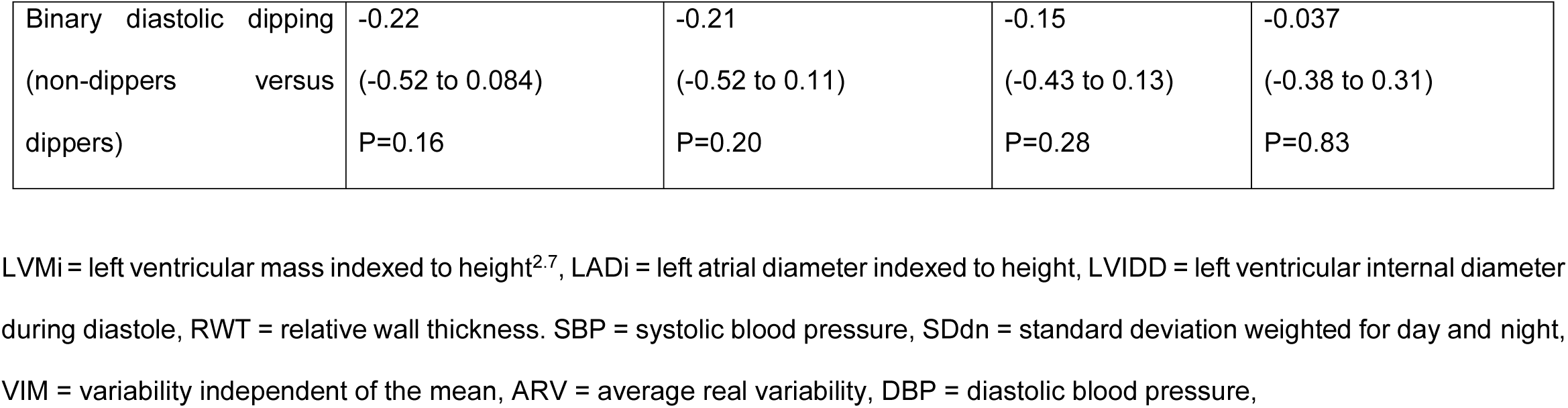
Associations of blood pressure measurements with cardiac structure, N=587. Analysis of multiply imputed data. Adjusted for sex, age at outcome assessment; maternal age at delivery, education, parity, and maternal pre-pregnancy BMI; household social class; smoking at age 17; minutes of moderate to vigorous physical activity at age 15; DXA-determined fat mass, height and height^2^ at age 17. Regression coefficients for continuous exposures are standardised, i.e. they represent the change in SDs of the outcome (cardiac structure measurement) per one SD higher blood pressure.

Clinic DBP was associated with higher RWT (β = 0.24, 95 % CI 0.15 to 0.33, P=1.4×10^−7^) and lower LADi and LVIDD. There was no evidence of an association between clinic DBP and LVMi^2.7^.

Results were broadly similar in the age and sex only adjusted models (Supplementary Table 4).

### Associations between ambulatory averages of BP and cardiac structures

There was evidence for a positive association between 24-hour mean SBP and LVMi^2.7^ (β = 0.17 SDs per SD higher 24-hour SBP, 95% CI 0.093 to 0.25, P=1.8×10^−5^), which was slightly smaller in magnitude than the association for clinic SBP (Figure 2). Daytime and night-time means for SBP also showed positive associations with LVMi^2.7^, with similar magnitudes to 24-hour mean SBP. The 24-hour mean SBP also showed a positive association with RWT (β = 0.18, 95% CI 0.089 to 0.26, P=8.1×10^−5^), with similar magnitudes of association seen for daytime and night-time mean SBP.

**Figure 2:**
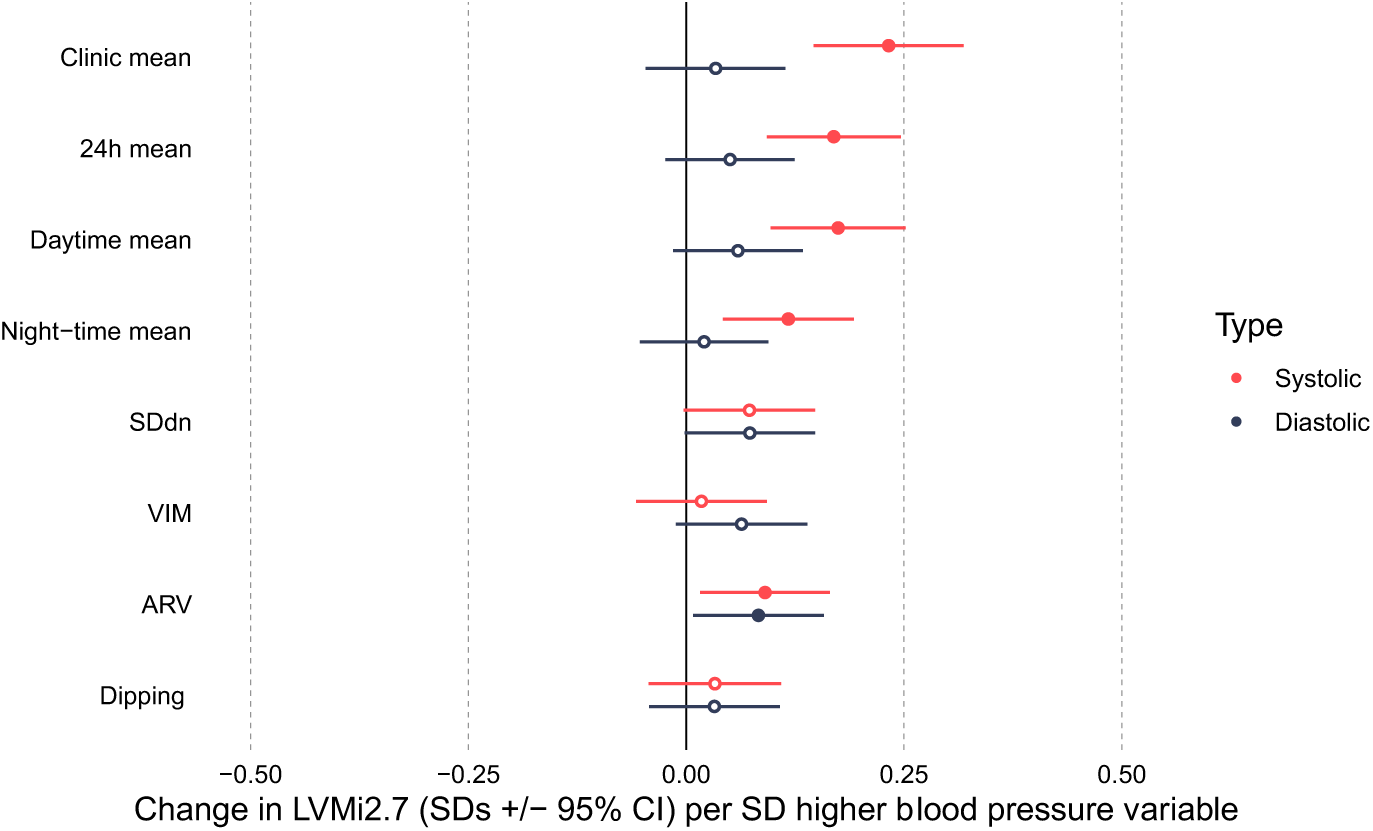
Forest plot of the mean difference in left ventricular mass indexed to height^2.7^ (LVMi^2.7^ in SDs +/- 95% confidence interval) per SD higher blood pressure (BP) variable in the confounder model. SDdn = Standard deviation weighted for day and night, VIM = variability independent of the mean, ARV = average real variability.

There was no evidence of associations between 24-hour, day-time or night-time mean SBP and LADi or LVIDD.

There was evidence for associations between all 24-hour DBP measures (mean, day and night) and RWT, with similar magnitudes of associations between the three exposures, but no evidence of associations for the other measures of cardiac structure.

### 2) Associations between 24-hour blood pressure variability and cardiac structures

ARV of SBP was associated with LVMi^2.7^ after adjustment for confounders (Table 2). All three measures of SBP variability (SDdn, ARV, VIM) were positively associated with RWT (SBP SDdn and RWT: β = 0.15, 95% CI 0.061 to 0.23, P=7.9×10^−4^). There was no consistent evidence of associations between SBP variability and LADi or LVIDD.

DBP variability measures were positively associated with RWT, although evidence of association was weaker for VIM than for SDdn and ARV. ARV and VIM of DBP were also positively associated with LVMi^2.7^ and LADi.

After further adjustment for 24-hour BP (Table 3), associations of SBP and DBP variability with LVMi^2.7^ and RWT attenuated towards the null. Some associations with RWT remained: before adjustment for mean DBP the standardised association between ARV of DBP and RWT was 0.13 (95% CI 0.045 to 0.21, P=2.7×10^−3^). After adjustment for mean DBP it was 0.11 (95% CI 0.022 to 0.19, P=0.014).

**Table 3.** Associations of BP variability and dipping with cardiac structure after adjustment for 24-hour mean BP, N=587. Analysis of multiply imputed data. Adjusted for sex, age at outcome assessment; maternal age at delivery, education, parity, and maternal pre-pregnancy BMI; household social class; smoking at age 17; minutes of moderate to vigorous physical activity at age 15; DXA-determined fat mass, height and height^2^ at age 17; mean 24-hour blood pressure (systolic or diastolic, as appropriate the exposure). Regression coefficients are standardised, i.e. they represent the change in SDs of the outcome (cardiac structure measurement) per one SD higher blood pressure.

| Exposure | Mean difference in cardiac structure measures (SDs) per SD higher BP: $\beta$, 95% confidence interval, P value | | | |
| --- | --- | --- | --- | --- |
|  | LVMi | LADi | LVIDD | RWT |
| SDdn SBP | 0.020<br>(-0.059 to 0.099)<br>P=0.62 | 0.024<br>(-0.060 to 0.11)<br>P=0.58 | -0.027<br>(-0.10 to 0.046)<br>P=0.47 | 0.10<br>(0.011 to 0.19)<br>P=0.028 |
| VIM of SBP | 0.020<br>(-0.054 to 0.094)<br>P=0.59 | 0.031<br>(-0.048 to 0.11)<br>P=0.44 | -0.022<br>(-0.091 to 0.047)<br>P=0.53 | 0.090<br>(0.0054 to 0.17)<br>P=0.037 |
| ARV of SBP | 0.044<br>(-0.034 to 0.12)<br>P=0.27 | 0.076<br>(-0.0065 to 0.16)<br>P=0.071 | 0.0060<br>(-0.066 to 0.078)<br>P=0.87 | 0.077<br>(-0.011 to 0.17)<br>P=0.085 |
| Systolic dipping (continuous) | 0.044<br>(-0.031 to 0.12)<br>P=0.35 | 0.038<br>(-0.042 to 0.12)<br>P=0.35 | 0.061<br>(-0.009 to 0.13)<br>P=0.086 | -0.057<br>(-0.14 to 0.028)<br>P=0.19 |
| Binary systolic dipping (non-dippers<br>versus dippers) | -0.14<br>(-0.31 to 0.041)<br>P=0.13 | -0.072<br>(-0.26 to 0.12)<br>P=0.45 | -0.043<br>(-0.21 to 0.12)<br>P=0.61 | -0.039<br>(-0.24 to 0.16)<br>P=0.71 |
| SDdn DBP | 0.066<br>(-0.01 to 0.14)<br>P=0.089 | 0.081<br>(0.0013 to 0.16)<br>P=0.046 | -0.012<br>(-0.082 to 0.058)<br>P=0.74 | 0.13<br>(0.41 to 0.21)<br>P=3.7x10<sup>-3</sup> |
| VIM of DBP | 0.076<br>(-0.0007 to 0.15)<br>P=0.052 | 0.093<br>(0.012 to 0.17)<br>P=0.024 | -0.0002<br>(-0.071 to 0.070)<br>P=0.99 | 0.12<br>(0.033 to 0.20)<br>P=6.9x10<sup>-3</sup> |
| ARV of DBP | 0.076<br>(-0.0007 to 0.15)<br>P=0.052 | 0.14<br>(0.055 to 0.21)<br>P=9.2x10<sup>-4</sup> | 0.0021<br>(-0.068 to 0.072)<br>P=0.95 | 0.11<br>(0.022 to 0.19)<br>P=0.014 |
| Diastolic dipping (continuous) | 0.031<br>(-0.044 to 0.11)<br>P=0.42 | 0.050<br>(-0.028 to 0.13)<br>P=0.21 | 0.034<br>(-0.034 to 0.10)<br>P=0.33 | -0.020<br>(-0.10 to 0.065)<br>P=0.65 |
| Binary diastolic dipping (non-dippers versus dippers) | -0.24<br>(-0.54 to 0.068)<br>P=0.13 | -0.18<br>(-0.50 to 0.13)<br>P=0.25 | -0.14<br>(-0.4 to 0.14)<br>P=0.33 | -0.076<br>(-0.42 to 0.27)<br>P=0.66 |

LVMi = left ventricular mass indexed to height<sup>2.7</sup>, LADi = left atrial diameter indexed to height, LVIDD = left ventricular internal diameter during diastole, RWT = relative wall thickness. SBP = systolic blood pressure, DBP = diastolic blood pressure, SDdn = standard deviation weighted for day and night, VIM = variability independent of the mean, ARV = average real variability

### 3) Associations between night-time BP dipping and cardiac structures

The results provided no evidence for associations between either of the dipping variables (percentage difference and categorical) and cardiac structures (Supplementary Table 4, Table 2, Table 3). This was true for both SBP and DBP.

### Complete case analysis

For all analyses, there were similar magnitudes of estimates between the complete cases and imputed analyses (Supplementary Table 2 and Table 3). However, as there was less power in the complete case analysis, confidence intervals were wider.

### Sensitivity analysis

A likelihood ratio test was performed to compare the association between average blood pressure (in quintiles as both a categorical and continuous variable) and blood pressure variability. Results suggest that this relationship is nonlinear (Supplementary Table 3). The same was performed for blood pressure/blood pressure variability and cardiac structure outcomes, which suggested that these associations are indeed linear (Supplementary Table 3). Regression models were then repeated for exposures related to blood pressure variability and dipping, but adjusting for categorical quintiles of blood pressure rather than continuous. Estimates from this analysis (Supplementary Table 5) were broadly similar to Table 3.

## Discussion

In this cross-sectional study of a general population of adolescents, we explored the association between both blood pressure variability and dipping and measures of cardiac structure. Average 24-hour and clinic blood pressure measurements showed positive associations of a similar magnitude with cardiac structures such as RWT, for both systolic and diastolic measures. Measures of 24-hour variability (SDdn and ARV) were positively associated with RWT, with ARV also showing a positive association with LVMi^2.7^. After adjustment for 24-hour mean BP, some associations persisted including the association between ARV of DBP and RWT. No associations were found between night-time dipping and cardiac structures in this cohort.

Variability in BP over 24 hours in this sample of adolescents was similar in magnitude to that reported in studies of adults [41, 42]. In contrast, the percentage of normal dipping was higher than in adult studies [28]. In adults, greater variability in BP and non-dipping are associated with cardiovascular risk, independently of average BP [2, 28, 43]. Two previous studies, restricted to hypertensive children, did not find an association between 24-hour BP variability and LVMi^2.7^ [16, 44]. Similarly, several studies have found little association between night-time dipping and LVMi in hypertensive children [27, 45, 46]. To our knowledge, this is the first study to explore these associations in a general population cohort of adolescents.

Higher mean SBP is associated with higher LVMi^2.7^ and RWT in our study. This finding, together with our previous finding that higher BMI is causally related to higher LV mass [14], suggests that higher values of LVMi^2.7^ and RWT are, on average, related to adverse cardiovascular health even in this young population, rather than due to high levels of fitness. This implies that the cardiac structures are meaningful markers of cardiac health in this young population. Both DBP and SBP were associated with RWT to a similar extent. However, unlike SBP, DBP did not show associations with LVMi^2.7^. This could reflect a greater importance of systolic pressure (and by implication pulse pressure on LV mass). It may also be at least partially driven by regression dilution bias [47], which is the biasing of the regression slope to towards zero, because of the greater levels measurement error for DBP compared with SBP [48].

Our results indicate that some associations between greater BP variability and cardiac structure remain once average BP was accounted for, such as the positive association between diastolic measure of ARV and RWT. These findings support the notion that the influence of BP variability on cardiac structure may begin early in life [49]. It is unclear which measurement used is superior in determining blood pressure variability as a risk factor. SDdn accounts for length of day and night, however it is still dependent on mean BP. The derivation of VIM attempts to overcome this issue as dividing the SD by the mean to the power *x* removes correlation between mean and SD [50]. ARV uses the differences in consecutive BP readings, therefore it is able to capture more frequent spikes in BP than the other methods [51]. Both systolic and diastolic ARV have been suggested to predict total and cardiovascular mortality [43]. ABPMs are currently recommended for diagnosis of hypertension among adolescents [52]; this study suggests that other ABPM-derived information such as ARV might aid identification of adolescents who are at an increased cardiovascular risk. In general, all three measures of blood pressure variability included provided similar magnitudes of associations with cardiac structures.

We found no convincing evidence for an association between non-dipping and cardiac structure in young people. Findings in older adults are inconsistent [53] and most studies finding a positive association between non-dipping and LV mass have been conducted in hypertensive individuals [54]. The majority of the participants in our sample had blood pressures in the normotensive range; other studies which included such participants have also not found evidence of an association [55].

The current study has several limitations. It is possible that our study may have lacked statistical power to detect some associations between BP variability and dipping and cardiac structure independently of mean BP. Furthermore, the cohort are of European descent and in a localised area of the UK, which may limit its generalisability. The study uses cross-sectional data, which limits our ability to determine the true direction of the association between blood pressure and cardiac structures, and whether this relationship may be causal. The participants included in our analysis are more affluent than the full ALSPAC cohort [19]. However, whilst this does affect the generalisability of the study, it does not necessarily lead to bias in the estimates of associations. ABPMs have been reported to affect sleep quality due to cuff inflation. This may affect night time dipping levels and therefore weaken associations [56]. Additionally, we were not able to assess longer term blood pressure variability, including visit-to-visit variability, which may be another meaningful value in adolescents to predict adult hypertension [49].

Our results show that, in adolescents, higher clinic and 24-hour BP, as well as an increase in blood pressure variability, are associated with more adverse cardiac structure. Non-dipping was not found to be associated with cardiac structure. Our study implies that measurement of BP variability, but not night-time dipping, might add to the assessment of cardiovascular risk in adolescents. However, this finding would benefit from replication in larger studies. It would be valuable to explore whether BP variability and dipping in adolescents track across the life course, and whether these BP measurements in adolescents are predictive of longer-term cardiovascular outcomes.

## Data Availability

Because of the sensitive nature of the data collected for this study, requests to access the dataset from qualified researchers trained in human subject confidentiality protocols may be submitted via the Avon Longitudinal Study of Parents and Children (ALSPAC) website http://www.bristol.ac.uk/alspac/researchers/access/.

## Acknowledgements

We are extremely grateful to all the families who took part in this study, the midwives for their help in recruiting them, and the whole ALSPAC team, which includes interviewers, computer and laboratory technicians, clerical workers, research scientists, volunteers, managers, receptionists and nurses. We thank Kirsten Leyland for her support with analyses.

## Supplementary materials

Supplementary file (Supplementary_Tables_Int_J_hyp.xlsx) contains four supplementary tables

1. **Supplementary Table 1a** : Distributions of imputed characteristics in the imputation datasets and in observed data (i.e. without imputation).
2. **Supplementary Table 1b**: Comparing participants included in the analysis with those excluded due to missing data
3. **Supplementary Table 2**. Complete case associations of BP variability and dipping with cardiac structure after adjustment for confounders and 24-hour mean BP
4. **Supplementary Table 3**. Likelihood ratio tests to assess non-linearity of associations
5. **Supplementary Table 4**. Associations of blood pressure measurements and 24-hour ambulatory blood pressure with cardiac structure (adjusted for age and sex)
6. **Supplementary Table 5**. Associations between blood pressure variability/dipping and cardiac structures adjusting for categorical quintiles of average blood pressure
7. **STROBE checklist**

## References

1. Rapsomaniki, E., et al., Blood pressure and incidence of twelve cardiovascular diseases: lifetime risks, healthy life-years lost, and age-specific associations in 1·25 million people. Lancet, 2014. 383(9932): p. 1899–911.

2. Yano, Y. and K. Kario, Nocturnal blood pressure and cardiovascular disease: a review of recent advances. Hypertens Res, 2012. 35(7): p. 695–701.

3. Hansen, T.W., et al., Predictive role of the nighttime blood pressure. Hypertension, 2011. 57(1): p. 3–10.

4. Wang, J., et al., Visit-to-visit blood pressure variability is a risk factor for allcause mortality and cardiovascular disease: a systematic review and meta-analysis. J Hypertens, 2017. 35(1): p. 10–17.

5. Stevens, S.L., et al., Blood pressure variability and cardiovascular disease: systematic review and meta-analysis. BMJ, 2016. 354: p. i4098.

6. de Swiet, M., P. Fayers, and E.A. Shinebourne, Blood pressure in first 10 years of life: the Brompton study. British Medical Journal, 1992. 304(6818): p. 23–26.

7. McCarron, P., et al., Blood pressure in young adulthood and mortality from cardiovascular disease. The Lancet, 2000. 355(9213): p. 1430–1431.

8. Levy, D., et al., Prognostic implications of echocardiographically determined left ventricular mass in the Framingham Heart Study. N Engl J Med, 1990. 322(22): p. 1561–6.

9. Armstrong, A.C., et al., Left atrial dimension and traditional cardiovascular risk factors predict 20-year clinical cardiovascular events in young healthy adults: the CARDIA study. Eur Heart J Cardiovasc Imaging, 2014. 15(8): p. 893–9.

10. Mancusi, C., et al., Left atrial dilatation: A target organ damage in young to middle-age hypertensive patients. The Campania Salute Network. Int J Cardiol, 2018. 265: p. 229–233.

11. Gaasch, W.H. and M.R. Zile, Left ventricular structural remodeling in health and disease: with special emphasis on volume, mass, and geometry. J Am Coll Cardiol, 2011. 58(17): p. 1733–40.

12. Wang, S., et al., Left ventricular hypertrophy, abnormal ventricular geometry and relative wall thickness are associated with increased risk of stroke in hypertensive patients among the Han Chinese. Hypertens Res, 2014. 37(9): p. 870–4.

13. Di Tullio, M.R., et al., Left ventricular mass and geometry and the risk of ischemic stroke. Stroke, 2003. 34(10): p. 2380–4.

14. Wade, K.H., et al., Assessing the causal role of body mass index on cardiovascular health in young adults: Mendelian randomization and recall-by-genotype analyses. Circulation, 2018. 138(20): p. 2187–2201.

15. Sega, R., et al., Blood pressure variability and organ damage in a general population: results from the PAMELA study (Pressioni Arteriose Monitorate E Loro Associazioni). Hypertension, 2002. 39(2 Pt 2): p. 710–4.

16. Bjelakovic, B., et al., Blood pressure variability and left ventricular mass index in children. J Clin Hypertens (Greenwich), 2013. 15(12): p. 905–9.

17. Zimpfer, M. and S.F. Vatner, Effects of acute increases in left ventricular preload on indices of myocardial function in conscious, unrestrained and intact, tranquilized baboons. J Clin Invest, 1981. 67(2): p. 430–8.

18. Cikes, M. and S.D. Solomon, Beyond ejection fraction: an integrative approach for assessment of cardiac structure and function in heart failure. Eur Heart J, 2016. 37(21): p. 1642–50.

19. Boyd, A., et al., Cohort Profile: the ‘children of the 90s’--the index offspring of the Avon Longitudinal Study of Parents and Children. Int J Epidemiol, 2013. 42(1): p. 111–27.

20. Fraser, A., et al., Cohort Profile: the Avon Longitudinal Study of Parents and Children: ALSPAC mothers cohort. Int J Epidemiol, 2013. 42(1): p. 97–110.

21. O’Brien, E., et al., European Society of Hypertension recommendations for conventional, ambulatory and home blood pressure measurement. J Hypertens, 2003. 21(5): p. 821–48.

22. Krause, T., et al., Management of hypertension: summary of NICE guidance. BMJ, 2011. 343: p. d4891.

23. Anstey, D.E., et al., Diagnosing Masked Hypertension Using Ambulatory Blood Pressure Monitoring, Home Blood Pressure Monitoring, or Both? Hypertension, 2018. 72(5): p. 1200–1207.

24. Bilo, G., et al., A new method for assessing 24-h blood pressure variability after excluding the contribution of nocturnal blood pressure fall. J Hypertens, 2007. 25(10): p. 2058–66.

25. Mena, L., et al., A reliable index for the prognostic significance of blood pressure variability. J Hypertens, 2005. 23(3): p. 505–11.

26. Hara, A., et al., Randomised double-blind comparison of placebo and active drugs for effects on risks associated with blood pressure variability in the Systolic Hypertension in Europe trial. PLoS One, 2014. 9(8): p. e103169.

27. Seeman, T., O. Hradský, and J. Gilík, Nocturnal blood pressure non-dipping is not associated with increased left ventricular mass index in hypertensive children without end-stage renal failure. Eur J Pediatr, 2016. 175(8): p. 1091–7.

28. Salles, G.F., et al., Prognostic Effect of the Nocturnal Blood Pressure Fall in Hypertensive Patients: The Ambulatory Blood Pressure Collaboration in Patients With Hypertension (ABC-H) Meta-Analysis. Hypertension, 2016. 67(4): p. 693–700.

29. Lang, R.M., et al., Recommendations for chamber quantification: a report from the American Society of Echocardiography’s Guidelines and Standards Committee and the Chamber Quantification Writing Group, developed in conjunction with the European Association of Echocardiography, a branch of the European Society of Cardiology. J Am Soc Echocardiogr, 2005. 18(12): p. 1440–63.

30. de Simone, G., et al., Left ventricular mass and body size in normotensive children and adults: assessment of allometric relations and impact of overweight. J Am Coll Cardiol, 1992. 20(5): p. 1251–60.

31. Banegas, J.R., et al., Relationship between Clinic and Ambulatory Blood-Pressure Measurements and Mortality. N Engl J Med, 2018. 378(16): p. 1509–1520.

32. Howe, L.D., D.A. Lawlor, and C. Propper, Trajectories of socioeconomic inequalities in health, behaviours and academic achievement across childhood and adolescence. J Epidemiol Community Health, 2013. 67(4): p. 358–64.

33. Laitinen, T.T., et al., Association of Socioeconomic Status in Childhood With Left Ventricular Structure and Diastolic Function in Adulthood: The Cardiovascular Risk in Young Finns Study. JAMA Pediatr, 2017. 171(8): p. 781–787.

34. Harville, E.W., J.W. Apolzan, and L.A. Bazzano, Maternal Pre-Pregnancy Cardiovascular Risk Factors and Offspring and Grandoffspring Health: Bogalusa Daughters. Int J Environ Res Public Health, 2018. 16(1).

35. Sterne, J.A. and G. Davey Smith, Sifting the evidence-what’s wrong with significance tests? BMJ, 2001. 322(7280): p. 226–31.

36. Amrhein, V., S. Greenland, and B. McShane, Scientists rise up against statistical significance. Nature, 2019. 567(7748): p. 305–307.

37. Perneger, T.V., What’s wrong with Bonferroni adjustments. BMJ, 1998. 316(7139): p. 1236–8.

38. Harrington, D., et al., New Guidelines for Statistical Reporting in the. N Engl J Med, 2019. 381(3): p. 285–286.

39. White, I.R., P. Royston, and A.M. Wood, Multiple imputation using chained equations: Issues and guidance for practice. Stat Med, 2011. 30(4): p. 377–99.

40. White, I.R. and J.B. Carlin, Bias and efficiency of multiple imputation compared with complete-case analysis for missing covariate values. Stat Med, 2010. 29(28): p. 2920–31.

41. Muntner, P., et al., Low correlation between visit-to-visit variability and 24-h variability of blood pressure. Hypertens Res, 2013. 36(11): p. 940–6.

42. Madden, J.M., et al., Correlation between short-term blood pressure variability and left-ventricular mass index: a meta-analysis. Hypertens Res, 2016. 39(3): p. 171–7.

43. Hansen, T.W., et al., Prognostic value of reading-to-reading blood pressure variability over 24 hours in 8938 subjects from 11 populations. Hypertension, 2010. 55(4): p. 1049–57.

44. Leisman, D., et al., Blood pressure variability in children with primary vs secondary hypertension. J Clin Hypertens (Greenwich), 2014. 16(6): p. 437–41.

45. Belsha, C.W., et al., Influence of diurnal blood pressure variations on target organ abnormalities in adolescents with mild essential hypertension. Am J Hypertens, 1998. 11(4 Pt 1): p. 410–7.

46. Lee, H., et al., Left ventricular hypertrophy and diastolic function in children and adolescents with essential hypertension. Clin Hypertens, 2015. 21: p. 21.

47. Hutcheon, J.A., A. Chiolero, and J.A. Hanley, Random measurement error and regression dilution bias. BMJ, 2010. 340: p. c2289.

48. Picone, D.S., et al., Accuracy of Cuff-Measured Blood Pressure: Systematic Reviews and Meta-Analyses. J Am Coll Cardiol, 2017. 70(5): p. 572–586.

49. Chen, W., et al., Adult hypertension is associated with blood pressure variability in childhood in blacks and whites: the bogalusa heart study. Am J Hypertens, 2011. 24(1): p. 77–82.

50. Mena, L.J., et al., 24-Hour Blood Pressure Variability Assessed by Average Real Variability: A Systematic Review and Meta-Analysis. J Am Heart Assoc, 2017. 6(10).

51. DeBarmore, B., et al., Association of ambulatory blood pressure variability with coronary artery calcium. J Clin Hypertens (Greenwich), 2018. 20(2): p. 289–296.

52. Flynn, J.T. and B.E. Falkner, New Clinical Practice Guideline for the Management of High Blood Pressure in Children and Adolescents. Hypertension, 2017. 70(4): p. 683–686.

53. Fagard, R., J.A. Staessen, and L. Thijs, The relationships between left ventricular mass and daytime and night-time blood pressures: a meta-analysis of comparative studies. J Hypertens, 1995. 13(8): p. 823–9.

54. Woodiwiss, A.J., et al., Impact of Blunted Nocturnal Blood Pressure Dipping on Cardiac Systolic Function in Community Participants Not Receiving Antihypertensive Therapy. Am J Hypertens, 2018. 31(9): p. 1002–1012.

55. Bello, N.A., et al., Associations of awake and asleep blood pressure and blood pressure dipping with abnormalities of cardiac structure: the Coronary Artery Risk Development in Young Adults study. J Hypertens, 2020. 38(1): p. 102–110.

56. Henskens, L.H., et al., Subjective sleep disturbance increases the nocturnal blood pressure level and attenuates the correlation with target-organ damage. J Hypertens, 2011. 29(2): p. 242–50.

